# Association between carotid arterial strain and heart rate variability in older age

**DOI:** 10.1101/2025.09.23.25336429

**Authors:** Matthew A Stanley, Massimiliano Fornasiero, Matt Webber, James C Moon, Peter Friberg, Alun D Hughes, Cristian Topriceanu, Gabriella Captur

## Abstract

**Background:** Loss of the normal heart rate variability (HRV) predicts death. We hypothesised that impaired arterial compliance and increased vascular stiffness, could inhibit the baroreceptor reflex, resulting in cardiac autonomic dysfunction and loss of the normal HRV.

**Aims:** To investigate the association between carotid arterial strain (CAS) and HRV in an older age population-based cohort.

**Method:** Participants (60-64 years) were from the 1946 Medical Research Council, National Survey of Health and Development British birth cohort. Carotid intima media thickness (cIMT) and CAS (exposures) were measured by ultrasound and time- and frequency-domain HRV indices (outcomes) by a resting 5-minute 12-lead tachogram. Generalized linear models were used, adjusted for relevant clinic-demographic confounders and subjected to sensitivity analysis in which we re-analyzed associations after additional adjustment for cIMT and after removing participants with known cardiovascular disease.

**Results:** 896 participants were included. On univariate analysis CAS was associated with HRV markers: standard deviation of normal-to-normal beats (SDNN), root mean square of successive differences (RMSDD), HRV triangular index, high- and low-frequency (H/LF) normalised high-frequency power (all *p*<0.05). Associations persisted in fully confounder-adjusted models: SDNN β=0.52 [confidence interval: 0.2,0.8] *p*<0.001; RMSDD β=0.59 [0.3,0.9], *p*<0.001; HRV triangular index β=-0.34 [-0.5,-0.1], *p*<0.001; HF power β=8.33 [2.2,14.4], *p*=0.007; LF power *β*=8.47 [1.0,15.9], *p*=0.026; normalised HF power β=0.55 [0.2,0.9], *p*=0.006. Key associations persisted in the sensitivity analysis.

**Conclusion:** Regardless of carotid atherosclerotic vascular disease (indicated by cIMT), hypertension or stroke, increased carotid arterial function in older age associates with a loss of HRV, potentially through an impaired baroreceptor response.

## Introduction

It has been suggested that reduced carotid arterial strain (CAS) caused by atherosclerosis, aging and metabolic factors such as hyperglycaemia^1^, reduces the sensitivity of carotid baroreceptors to blood pressure changes^2^ resulting in cardiac autonomic dysfunction. Such autonomic dysfunction is known to manifest amongst other things, as a dampening of heart rate variability (HRV). CAS ultimately relates to the quantity and integrity of elastin, and organisation of collagen in the arterial wall. Indices of HRV are clinically obtained through time- and frequency-domain analysis of electrocardiogram (ECG) traces. Low HRV has been implicated in increased mortality, e.g. following myocardial infarction (MI)^3^ and in heart failure^4^.

An association between carotid stiffening and HRV has been found in young patients with type 1 diabetes^5^, and in young persons free from cardiovascular disease^6^, but whether this association persists into older age, at the population level is not known. We sought to investigate whether CAS and carotid intima-media thickness (cIMT) were associated with HRV in an older age British-based cohort.

## Methods

### Participants

Participants were from the Medical Research Council (MRC) National Survey of Health and Development (NSHD)–a birth cohort study comprised of 5,362 individuals born in 1 week in 1946 in Britain. The cohort has been extensively followed up with periodic assessments which have been described elsewhere^7^. Briefly, the cohort has been evaluated multi-dimensionally: anthropometrically, socio-economically, and in terms of life-style choices (e.g., smoking) and health function (e.g., mental health, cardiovascular and respiratory function)^7^. The current cross-sectional study uses data collected between 2006-2010 when participants were aged 60-64 years of age. Written, informed consent was obtained from all participants and ethical approval was granted from the Greater Manchester Local Research Ethics Committee and the Scotland Research Ethics Committee^7^. All procedures were in accordance with the ethical standards of our institutional and/or national research ethics committees and conformed to the 1964 Helsinki declaration and its later amendments or comparable ethical standards.

The participant selection process is shown in **Figure 1**. The minimum set of inclusion criteria comprised the availability of bilateral CD ultrasonic data and standard deviation of normal-to-normal beats (SDNN) HRV data. Participants were sent a postal invite and pre-assessment questionnaire. The questionnaire collected data on socio-demographic factors, lifestyle and medical history.

**Figure 1.**
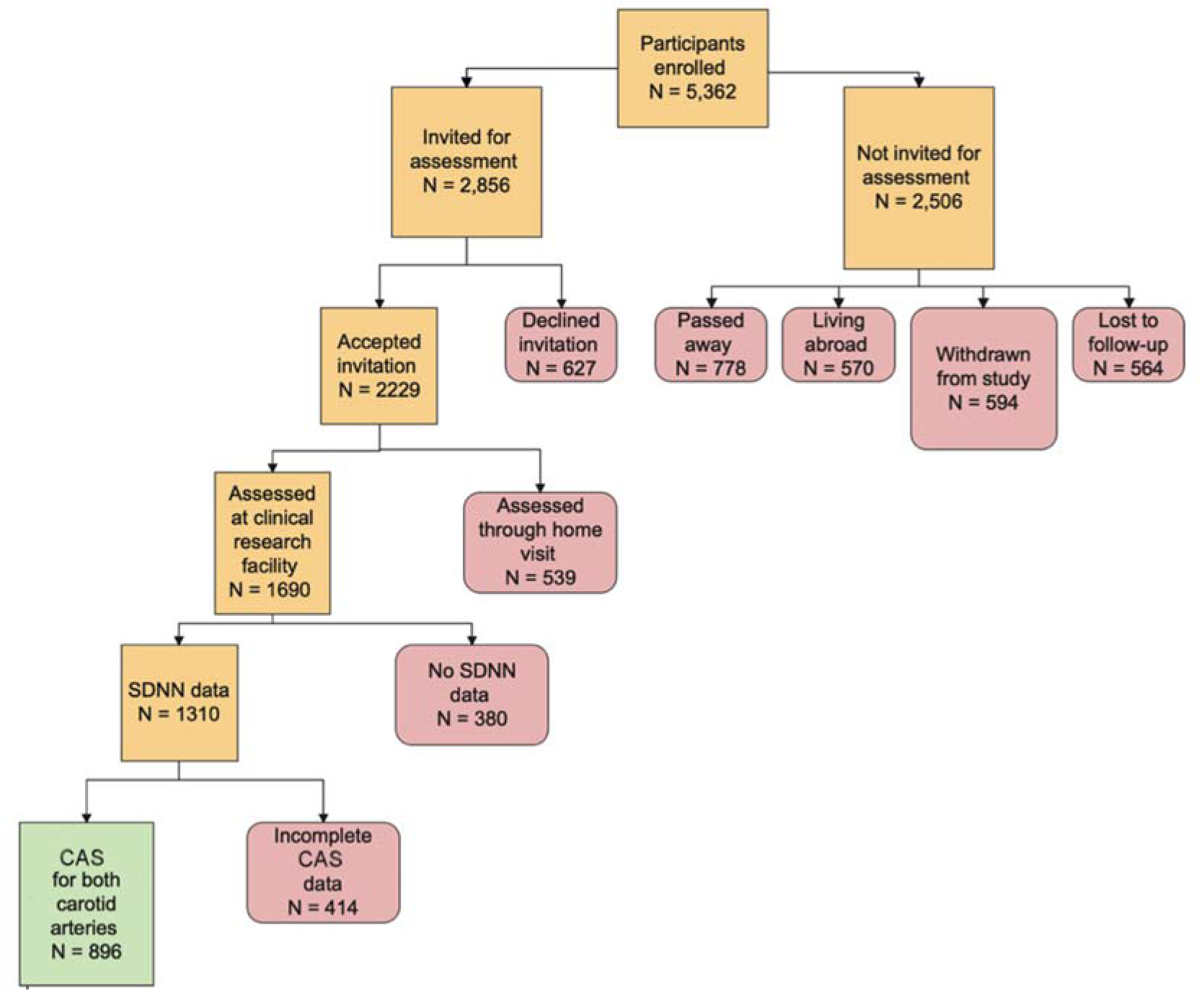
Flowchart summarising participant inclusion. Loss to follow-up over time was a concern in NSHD. Those with lower educational attainment, lower childhood cognition and lifelong smokers were less likely to attend the 60-64 year assessment but the sample remained representative of the general population as per the 2001 UK census^50^. *NSHD = National Survey of Health and Development; SDNN = standard deviation of normal-to-normal beats*.

### Outcome: HRV at 60-64 years

HRV was assessed through a 5-minute, 12-lead ECG of the supine, rested participant. The ECG programme was specifically developed for HRV analysis by a member of the data-gathering team. Recordings were manually cleaned by a physician to remove artefacts, ensure that normal beats were all registered and that ectopics were discarded. The analysis was completed automatically by CardioNavigator Plus (Spacelabs Healthcare Ltd, Snoqualmie, Washington) to generate values for the following HRV parameters: SDNN, root mean square of successive differences (RMSDD) and high-frequency (HF) power as measures of parasympathetic activity; low-frequency (LF) power as a measure of sympathetic activity^8^. Additional indices also included normalised LF, normalised HF power, LF/HF ratio, HRV triangular index, total power spectral density (total PSD) and power spectral density squared.

### Exposures

CAS and cIMT were measured at the clinic visit with a GE Vivid I ultrasound scanner (GE Healthcare; Chalfont St Giles, UK) with a high-resolution probe (12Hz). Clear images of the artery, 1cm proximal to the bifurcation, were obtained. Ten second cineloops were recorded in digital imaging and communications in medicine format and downloaded for offline analysis by the Vascular Physiology Unit, Institute of Cardiovascular Science, University College London, using dedicated software (Carotid Analyser; Iowa City, Iowa). Images were calibrated and software used to automatically generate diameter measurements. Area strain (given as a percentage) was calculated as the difference between maximum and minimum cross-sectional area, as a proportion of the minimum cross-sectional area. Average strain was calculated as the mean of left and right strain. cIMT was calculated as previously described^9^.

### Covariates

Covariates were selected *a priori* based on their previously published association with HRV and added into our models successively, after centering on age, to help with the interpretation of coefficients. Model 1 adjusted for sex; Model 2 included additional adjustments for SEP; Model 3 added clinical covariates known to be associated with HRV; and Model 4 added cardiac covariates known to be associated with HRV. The same models were used for all the HRV outcomes.

The sex of participants was recorded as male or female (1/2). Height and weight measurements were taken in light, indoor clothing without shoes. Height was measured to the nearest millimetre using a portable stadiometer with the head in the Frankfort plane. Weight measurements to the nearest 0.1kg, were taken to calculate body mass index (BMI). Waist circumference measurements were taken at the midpoint between the costal margin and the iliac crest and hip circumference was measured at the level of the greater trochanter. The waist-to-hip ratio (WHR) was subsequently derived. Participants’ socio-economic position (SEP) was evaluated using occupational data from 1989, when they were in active working age, according to the UK Office of Population Censuses and Surveys Registrar General’s social class, dichotomized as manual or non-manual (0/1). Brachial systolic and diastolic blood pressure measurements were taken twice with the participant in a seated position using an Omron HEM-705 sphygmomanometer (OMRON UK Healthcare UK Ltd.; Milton Keynes, UK). The second reading (or the first if the second was missing) was used in our analysis. Two-dimensional transthoracic echocardiography was performed to measure left ventricular ejection fraction and mass as previously described^10^.

Information about medication usage relevant to HRV, was collected through survey instruments and self-reporting along with other relevant clinical information to capture history of diabetes, heart disease (i.e. ischemic heart disease, myocardial infarction, stroke, heart failure, heart rhythm abnormality, congenital heart disease, rheumatic heart disease, and other cardiovascular diseases), hypertension, physical activity (as self-reported activity in average minutes/day spent at a metabolic equivalent task of 1.5-2.99 in the last year) and smoking as previously described^11–13^.

For biochemical analysis, a 50ml blood sample was collected in clinic using the Sarstedt system (Sarstedt; Nümbrecht, Germany). Total cholesterol, high-density lipoprotein (HDL) cholesterol and triglyceride were measured using a Siemens Dimension Xpand analyser (Siemens plc Healthcare Sector; Frimley, UK) using the manufacturer’s assays. HbA1c was analysed using a TOSOH G7 analyser (Tosoh Bioscience Ltd; Redditch, UK). Low-density lipoprotein (LDL) was calculated by the Friedewald equation. A participant was defined as hypercholesterolaemic if LDL > 4.9mmol/L based on guidance from the National Institute of Health and Care Excellence^14^.

### Statistical analysis

Statistical analyses were conducted using R version 3.6.2 (RStudio Team 2020). Distribution of data was assessed using Q-Q plots, histograms and the Shapiro-Wilk test. Continuous sample variables are expressed as mean ± 1 standard deviation (SD) or median (interquartile range) as appropriate; categorical sample variables, as counts and percent. Differences between groups were tested using analysis of variance (ANOVA) with post-hoc Tukey test or else Kruskal-Wallis with post-hoc Nemenyi test for normally and non-normally distributed continuous data respectively, or with a Chi-square test for categorical data.

Due to the skewed distribution of HRV parameters, generalized linear models (GLMs) with a gamma distribution and log link were fitted to examine the associations of CD and cIMT with HRV. Unless otherwise stated, model coefficients (β) in results are expressed as percent change per unit increase in exposure (%Δ per unit), calculated as 100×[exp(β)−1]%. To determine whether the associations of CAS with HRV differed by sex, an interaction term for sex were tested at the 10% significance level and no interaction was found to justify stratification by sex. Where more than one measure of CAS was significant at univariate analysis, average CAS was used in the multivariable model. Model assumptions were verified with regression diagnostics. Multi-collinearity between final model variables was excluded by demonstrating variance inflation factors <3. Data missingness was minimal in the study sample (**Supplementary Table S1**) so multiple imputation was not required. Strength of evidence for an association was assessed on the basis of the size of the regression coefficients, their confidence interval (CI) and the *p* value. All tests were 2 sided.

We ran sensitivity analyses in which we re-analyzed the association between CAS and HRV biomarkers after removing participants with known cardiovascular disease and after additional adjustment for cIMT (**Supplementary Tables S2-S3**)

## Results

### Participant characteristics

Of the 5362 originally enrolled into NSHD, 747 were deceased, 570 had emigrated, 853 had withdrawn and 530 were not contactable, leaving 2662 that were successfully interviewed between 2006-2010. Of these 896 had contemporaneous 5-minute ECG for SDNN HRV (outcome) and carotid ultrasonography for CAS (exposure, **Figure 1**). Characteristics of study participants are presented in **Table 1**. The population mean for SDNN was 30.0 (IQR 23.1-39.6) with 46.5% being male. Participants with dampened HRV (lowest SDNN quartile) were more likely to be older, male and smokers, suffering from hypertension, cardiovascular disease, diabetes or hypercholesterolaemia. Data missingness for key covariates used in multivariable models per exposure-outcome pair are presented in **Supplementary Table S1**.

**Table 1.**
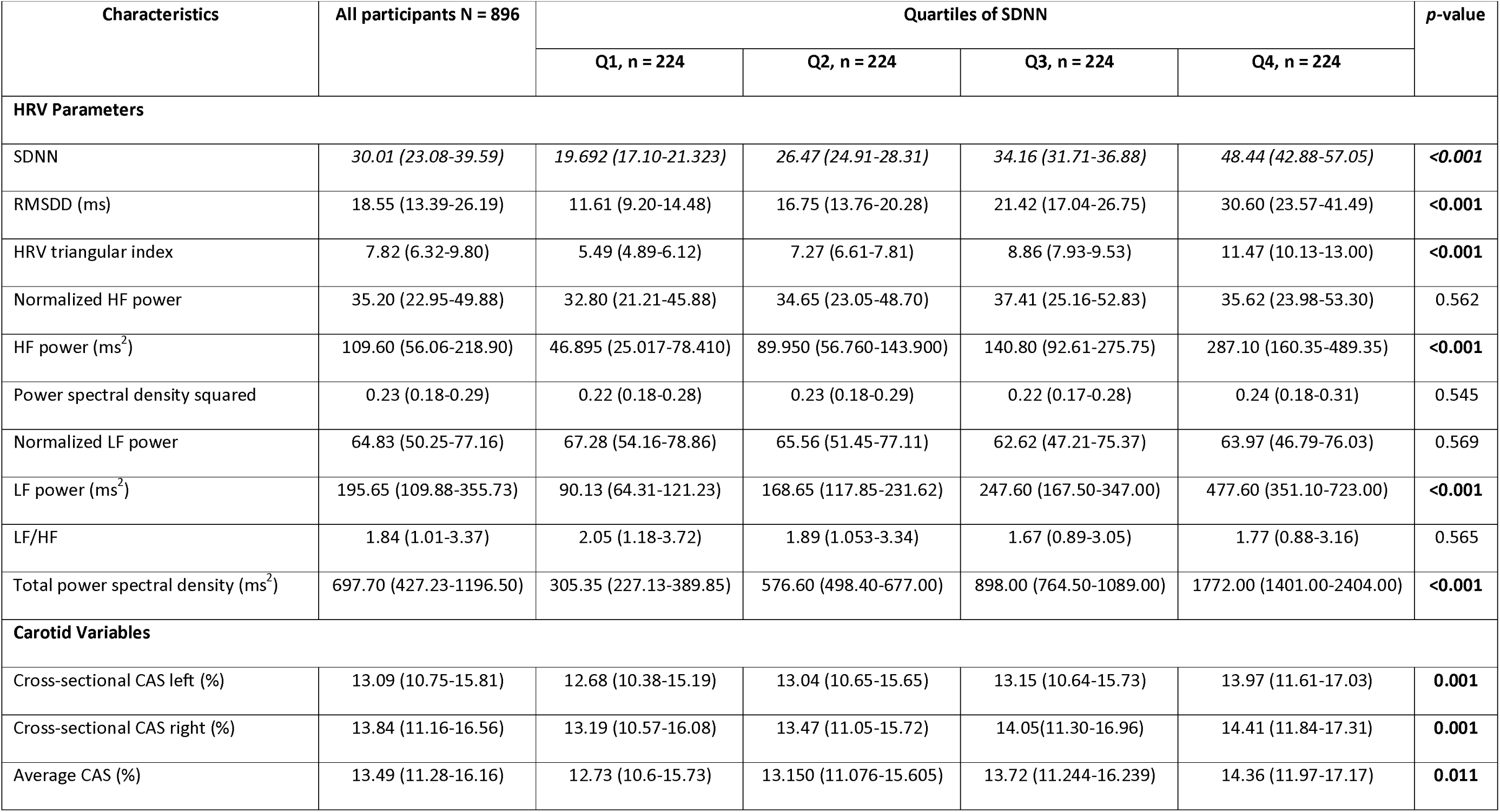
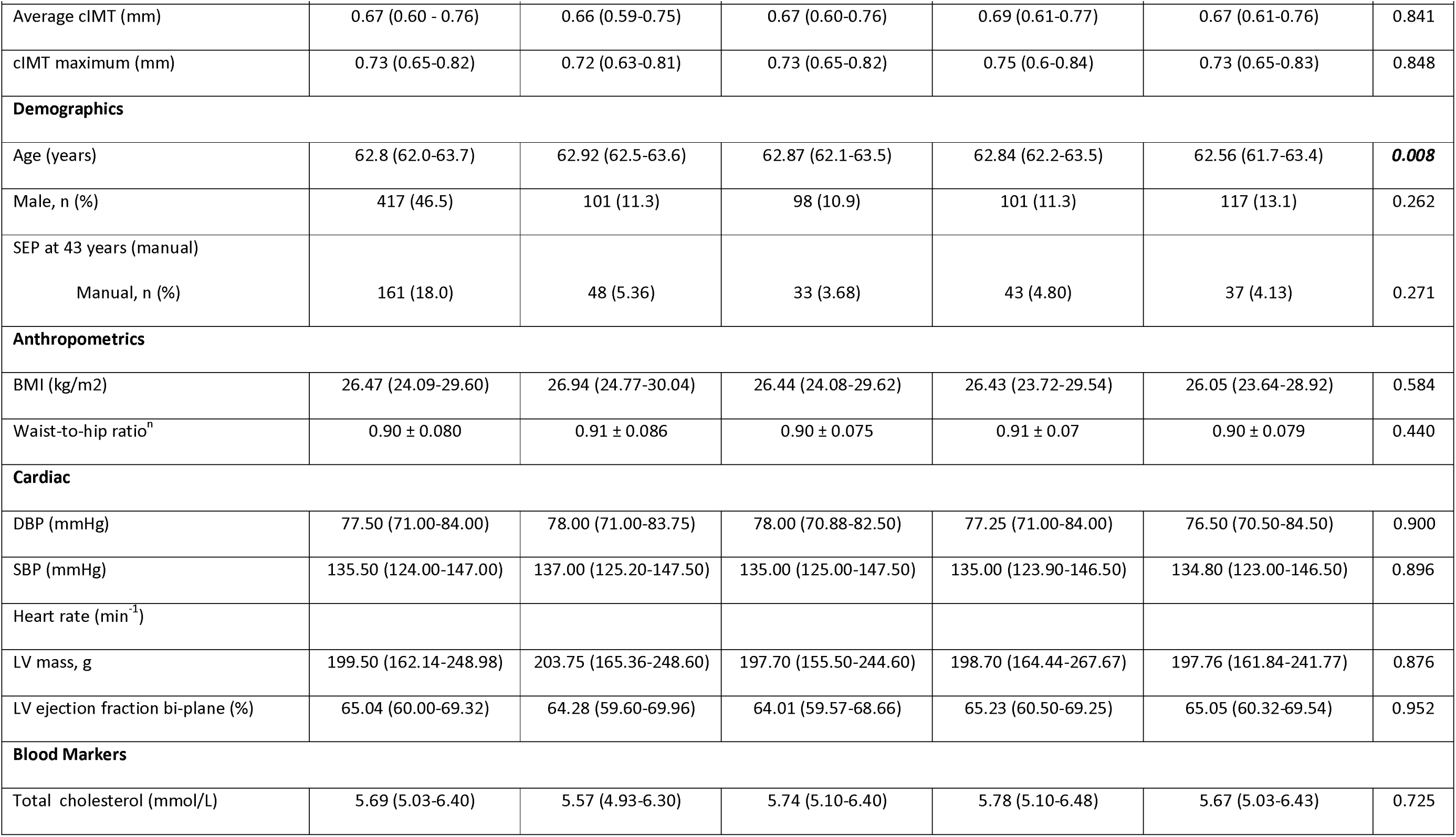
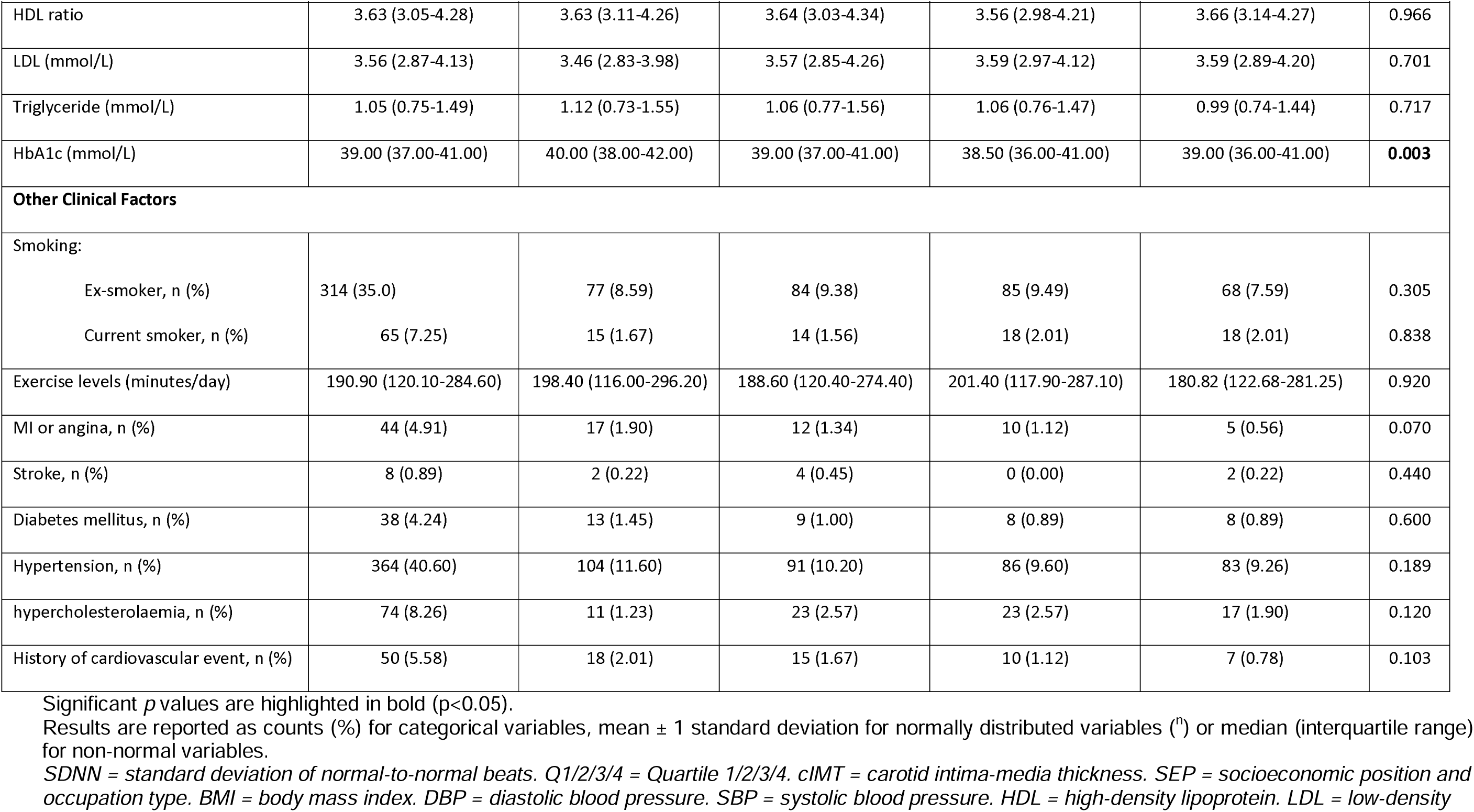
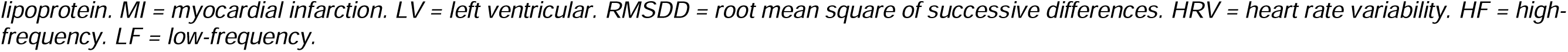
Clinicodemographic characteristics of the cohort according to SDNN quartiles.

### Associations of CAS with SDNN, RMSDD and HRV triangular index

Left (%Δ per unit=61.6%, 95% confidence interval [35.0% to 101.0% per unit], *p*<0.001), right (%Δ per unit=46.3% [22.1% to 82.2%], *p*<0.001) and average (%Δ per unit=69.8% [35.0% to 122.5%], *p*<0.001) cross-sectional CAS showed significant positive associations with SDNN on univariate analysis (**Table 2**), while age (%Δ per unit=–79.0% [–90.0% to –55.1%], *p*<0.001), sex (%Δ per unit=–84.0% [–97.3% to 0.0%], *p*=0.047), BMI (%Δ per unit=–20.6% [–33.0% to 0.0%], *p*=0.025), triglyceride levels (%Δ per unit=–76.3% [–90.0% to –33.0%], *p*=0.008), HbA1c (%Δ per unit=−18.1% [−25.9% to −9.5%], *p*<0.001) and previous MI or angina (%Δ per unit=−98.3% [−99.9% to −9.5%], *p*=0.029) showed significant negative associations with SDNN. In fully adjusted multivariable models, average CAS (%Δ per unit=68.2% [22.1% to 122.6%], *p*<0.001), age (%Δ per unit=−67.1% [−86.5% to −18.1%], *p*=0.016) and male sex (%Δ per unit=−93.6% [−99.3% to −45.1%], *p*=0.011) retained independent associations with SDNN (**Table 3**).

**Table 2.**
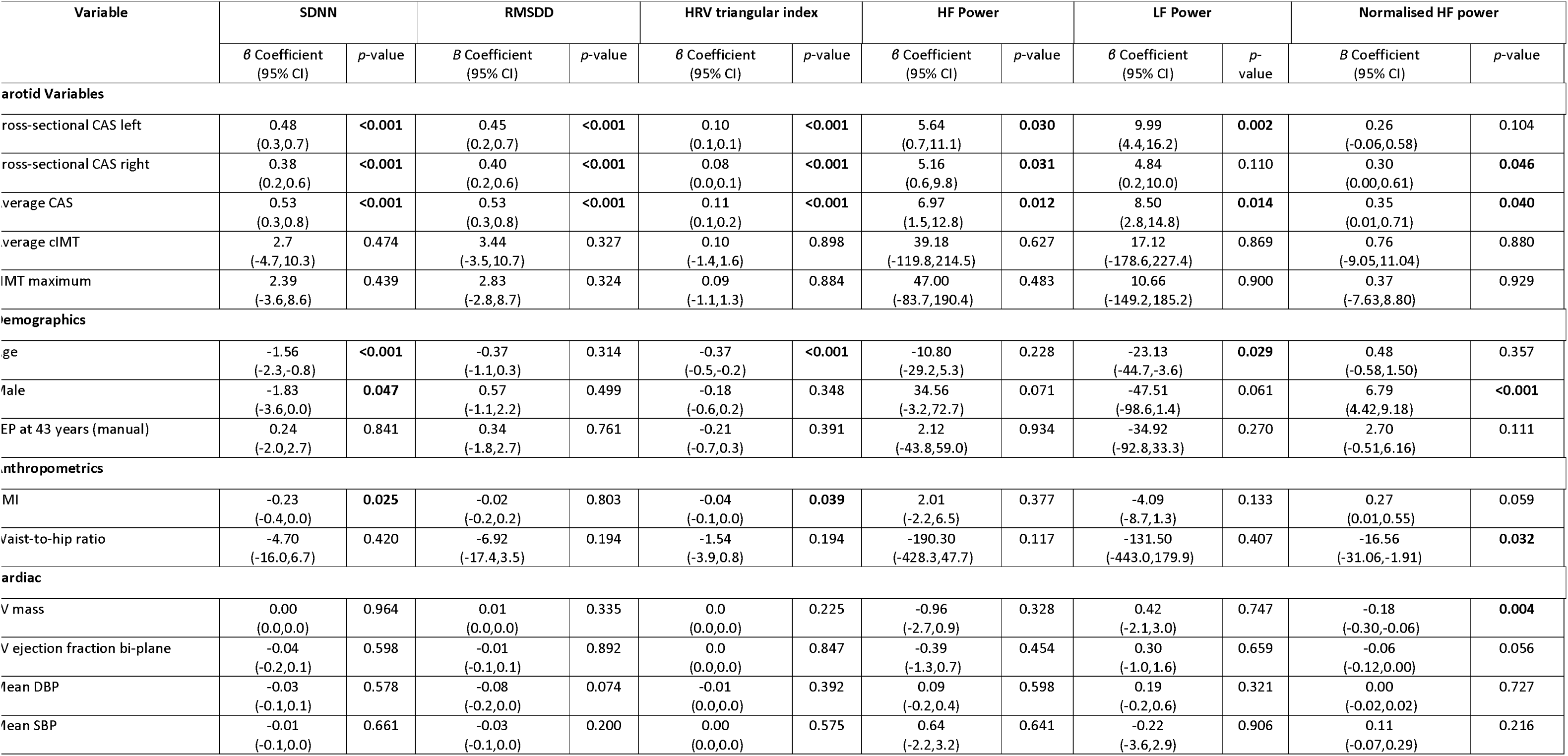
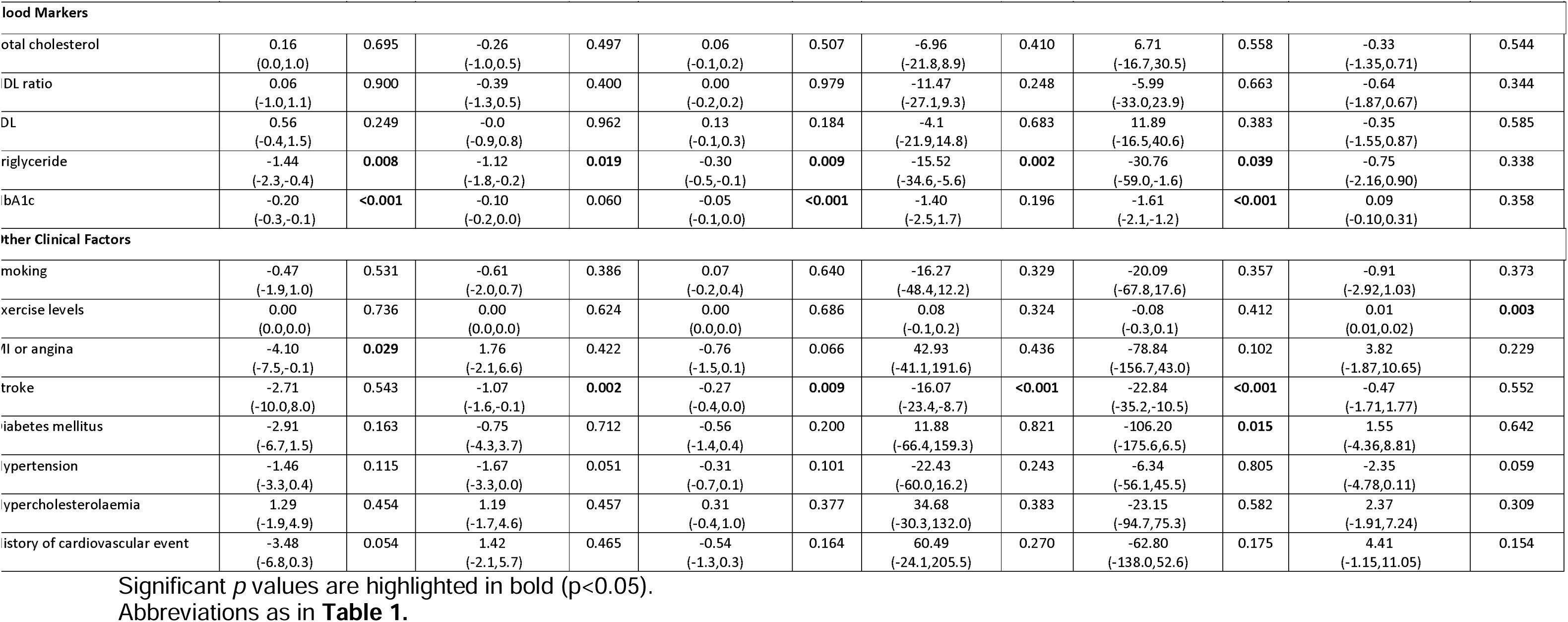
Univariate regression analysis for each exposure or covariate with HRV indices.

**Table 3.** Multivariable regression analysis for CD with HRV indices.

| Variable | SDNN |  | RMSDD |  | HRV Triangular Index |  | HF power |  | LF power |  | Normalised HF Power |  |
| --- | --- | --- | --- | --- | --- | --- | --- | --- | --- | --- | --- | --- |
| | $\beta$ Coefficient (95% CI) | *p*-value | $\beta$ Coefficient (95% CI) | *p*-value | $\beta$ Coefficient (95% CI) | *p*-value | $\beta$ Coefficient (95% CI) | *p*-value | $\beta$ Coefficient (95% CI) | *p*-value | $\beta$ Coefficient (95% CI) | *p*-value |
| age | -1.11<br>(-2.0,-0.2) | **0.016** | -0.12<br>(-1.0,0.7) | 0.781 | -0.34<br>(-0.5,-0.1) | **<0.001** | -6.23<br>(-23.8,11.3) | 0.486 | -8.17<br>(-31.8,15.4) | 0.497 | 0.39<br>(-0.8,1.5) | 0.507 |
| sex | -2.75<br>(-4.9,-0.6) | **0.011** | 0.44<br>(-1.6,2.5) | 0.670 | -0.34<br>(-0.8,0.1) | 0.149 | 34.98<br>(-7.6,77.5) | 0.107 | -62.80<br>(-117.6,-7.9) | **0.025** | 7.61<br>(4.8,10.4) | **<0.001** |
| average CAS | 0.52<br>(0.2,0.8) | **0.001** | 0.59<br>(0.3,0.9) | **<0.001** | 0.09<br>(0.0,0.2) | **0.005** | 8.33<br>(2.2,14.4) | **0.007** | 8.47<br>(1.0,15.9) | **0.026** | 0.55<br>(0.2,0.9) | **0.006** |
| EP at 43 years | -0.10<br>(-2.7,2.6) | 0.942 | -1.59<br>(-3.9,1.0) | 0.198 | -0.26<br>(-0.8,0.3) | 0.375 | -28.50<br>(-74.9,17.9) | 0.228 | -18.43<br>(-80.7,43.9) | 0.562 | -0.46<br>(-3.9,3.3) | 0.800 |
| MI | -0.06<br>(-0.3,0.2) | 0.646 | 0.05<br>(-0.2,0.3) | 0.730 | -0.02<br>(-0.1,0.0) | 0.486 | 2.56<br>(-3.1,8.2) | 0.375 | -4.06<br>(-10.2,2.1) | 0.194 | 0.11<br>(-0.3,0.5) | 0.577 |
| triglyceride | -0.94<br>(-2.4,0.6) | 0.207 | -1.10<br>(-2.4,0.4) | 0.109 | -0.19<br>(-0.5,0.2) | 0.252 | -19.69<br>(-43.1,3.7) | 0.098 | -6.61<br>(-38.3,25.0) | 0.682 | -0.72<br>(-2.6,1.4) | 0.466 |
| HbA1c | -0.10<br>(-0.2,0.0) | 0.143 | -0.03<br>(-0.1,0.1) | 0.634 | -0.03<br>(-0.1,0.0) | 0.061 | -0.18<br>(-2.9,2.6) | 0.900 | -1.06<br>(-3.3,1.2) | 0.350 | 0.08<br>(-0.1,0.3) | 0.461 |
| MI/angina | -2.46<br>(-6.8,2.7) | 0.302 | 2.81<br>(-2.0,9.0) | 0.298 | -0.37<br>(-1.4,0.8) | 0.485 | 107.56<br>(-52.2,267.4) | 0.187 | 26.62<br>(-78.0,131.3) | 0.618 | 5.92<br>(-1.1,14.6) | 0.134 |
| stroke | -0.89<br>(-1.8,0.7) | 0.142 | -0.68<br>(-1.5,1.2) | 0.230 | -0.14<br>(-0.4,0.2) | 0.339 | -9.18<br>(-26.6,8.2) | 0.300 | -15.55<br>(-35.8,4.7) | 0.132 | -0.62<br>(-2.0,2.1) | 0.524 |
| hypertension | -1.59<br>(-3.8,0.6) | 0.157 | -1.33<br>(-3.4,0.8) | 0.217 | -0.34<br>(-0.8,0.2) | 0.175 | -13.43<br>(-57.2,30.3) | 0.546 | -36.02<br>(-91.1,19.1) | 0.200 | -1.43<br>(-4.4,1.6) | 0.341 |

Only fully adjusted data for the final model (Model 4) are shown. Model 1 (centered on age) adjusted for sex; Model 2 additionally adjusted for SEP; Model 3 additionally adjusted for clinical covariates namely BMI, HbA1c and Triglycerides; Model 4 additionally adjusted for cardiac covariates namely, hypertension, prior stroke and previous MI or angina. Significant *p* values are highlighted in bold (*p*<0.05).
Abbreviations as in **Table 1**.

On univariate analysis, average, left and right CAS respectively, were all positively associated with RMSDD (%Δ per unit=69.8% [35.0% to 122.6%], *p*<0.001; =56.8% [22.1% to 101.4%], *p*<0.001; =49.2% [22.1% to 82.2%], *p*<0.001;). Triglyceride and stroke respectively were negatively associated with RMSDD (%Δ per unit=−67.4% [−83.5% to −18.1%], *p*=0.019; =−65.7% [−79.8% to −9.5%], *p*=0.002) (**Table 2**). In fully adjusted multivariable models only average CAS retained an independent association with RMSDD (%Δ per unit=80.4% [35.0% to 146.0%], *p*<0.001, **Table 3**). On univariate analysis, average, left and right CAS respectively, were all positively associated with HRV triangular index (%Δ per unit=11.6% [10.5% to 22.1%], *p*<0.001; =10.5% [10.5% to 10.5%], *p*<0.001; =8.3% [0.0% to 10.5%], *p*=<0.001, **Table 2**). Age (%Δ per unit=-30.9% [-39.4% to -18.1%], *p*<0.001), BMI (%Δ per unit=-25.9% [-39.4% to -9.5%], *p*=0.039), triglyceride (%Δ per unit=-25.9% [-39.4% to -9.5%], *p*=0.009), HbA1c (%Δ per unit=-4.9% [-9.5% to 0.0%], *p*<0.001) and stroke (%Δ per unit=-23.7% [-33.0% to 0.0%], *p*=0.009) were negatively associated with HRV triangular index. In fully adjusted multivariable models, only age and average CAS retained independent associations (%Δ per unit=-28.8% [-39.4% to - 9.5%], *p*<0.001; =9.42% [0.0% to 22.1%], *p*=0.005 **Table 3**)

### Associations of CAS with HF power, LF power and normalised HF power

Average, left and right CAS showed a significant positive association with HF power at univariate analysis (respectively %Δ per unit=10.6x10^4^% [3.5X10^2^% to 36.2x10^6^%], *p*=0.012; =28.0x10^3^% [101% to 66.2x10^5^%], *p*=0.031; =17.3x10^3^% [82.2% to 180.3x10^4^%], *p*=0.030, **Table 2**). On multivariable analysis, average CAS (%Δ per unit=41.5x10^4^% [802% to 17.9x10^7^%], *p*=0.007) retained a significant association (**Table 3**).

Average (%Δ per unit=49.2x10^4^% [15.4x10^2^% to 26.8x10^7^%], *p*<0.014) and left (%Δ per unit=21.8x10^5^% [8.0x10^3^%,10.8x10^8^%], *p*=0.002) CAS were significantly associated with LF power on univariate analysis. HbA1c negatively associated with LF power (%Δ per unit=-80% [-87.8% to -69.9%], *p*<0.001), as were age, triglyceride blood levels, previous stroke and diagnosis of diabetes mellitus (**Table 2**). On multivariable analysis, average CAS (%Δ per unit=47.7% [17.1x10^1^%,80.4x10^7^%], *p*=0.026) was significantly associated with LF power (**Table 3**).

Average and right CAS were associated with normalised HF power at univariate analysis (%Δ per unit=41.9% [1.0% to 103.4%], *p*=0.040; =35.0% [0.0% to 84.0%], *p*=0.046). Other associations are summarised in **Table 2**. Average CAS and sex retained significance in fully adjusted multivariable models (%Δ per unit=73.3% [22.1% to 146.0%], *p*=0.006; =20.2x10^4^% [12.1x10^3^% to 32.9x10^5^%], *p*<0.001; **Table 3**).

The association between average CAS with total power spectral density and PSD squared, was significant at univariate analysis but attenuated after multivariable adjustment (**Supplementary Table S4-S5**). There was no association between normalized LF power and LF/HF ratio with average CAS at univariate analysis.

### Sensitivity analysis

When removing participants with known cardiovascular disease from the analysis, average CAS retained association with SDNN, RMSDD and HRV triangular index (%Δ per unit=53.7% [10.5% to 101.4%], *p*=0.003; =63.2% [22.1% to 122.6%], *p*<0.001; =7.25% [0.0% to 10.5%], *p*=0.021, **Table S2**) and the same was observed after adjusting for cIMT (**Table S3**).

## Discussion

In a cross-sectional population-based study we found that older persons with stiffer carotids exhibited impairment of normal HRV.

Our study data show an independent association between CAS and several HRV measures including SDNN, RMSDD, HRV triangular index, HF/LF power, and normalised HF power. Results lend credence to our initial theory that reduced CAS could potentially dampen the sensitivity of carotid sinus baroreceptors thus reducing HRV. This is also consistent with previous studies reporting similar associations in younger cohorts^15,16^, healthy adults^17^, and in patients with hypertension^18^ and people with type 2 diabetes mellitus^19^.

Carotid vascular stiffness indices are significantly associated with endothelial dysfunction as measured by flow-mediated dilatation^20^. Given that endothelial changes precede atherosclerosis and correlate with disease severity in both early and late stages, carotid CAS may be a more sensitive atherosclerosis biomarker than cIMT. There is little evidence to support the role of lipid-lowering interventions on CAS, however, a ketogenic diet that increases LDL, has been shown to associate with a decrease in carotid distensibility (though not in cIMT^22^) in patients with difficult to treat epilepsy. Therefore, CAS may better reflect short-term changes in atherosclerotic risk factors compared to cIMT.

The majority of HRV parameters we appraised in this study showed association with both left and right CAS, with the sole exception of normalised HF power. The asymmetrical cardiac reflex response has long been recognized but data is conflicting with some studies that focused on the RR interval, reporting greater dependence on right carotid sinus stimulation^23, 24^, while another study found no left-right differences in the carotid-cardiac reflex responses^25^. The fairly consistent asymmetry identified in our study, could be related to differences in right/left-sided cardiac innervation and to different projections of baroreceptor afferents to the solitary tract nucleus^23^. Because stimulation of the right carotid sinus in various clinical scenarios may have a larger influence on RR interval variability, this may confound the association with HRV biomarkers for a given value of right CAS compared to the left, resulting in a stronger association for the left than the right, as seen in our study.

We found that although LF and HF power were associated with CAS, the LF/HF ratio was not. This is in agreement with another study assessing HRV and carotid vascular stiffness indices in hypertensive patients^18^. While LF and HF power increase as CAS increases, the rate of increase is such that the ratio between the two remains unchanged. LF and HF power were previously thought to reflect sympathetic and parasympathetic tone respectively^26^ but this view has been fairly strongly criticised. Our results would suggest that the reduced CAS affects both systems equally, so sympathovagal balance is maintained. This is contentious however, and it is likely that there is not such a well-defined boundary between representation of sympathetic and parasympathetic tone in HRV analysis. Recent studies suggest that LF power may be reflecting cardiac autonomic outflow by baroreflexes rather than true sympathetic tone^27,28^, in which case LF and HF power may be capturing non-distinct determinants of HRV, detracting from our ability to infer the sympathovagal balance.

We found a significant association between CAS and HRV but not between cIMT and HRV, despite both CAS and cIMT being putative biomarkers of carotid atherosclerotic severity. The published literature is similarly divided with some previous studies describing a significant inverse relationship between cIMT and HRV^29, 30^ and others finding no significant association^31,32^. Discrepant results could be attributed to study design differences with some factors such as participants’ age, physical activity levels and other clinically-relevant covariates not being adjusted for where associations were reported. The baroreflex is initiated in response to baroreceptor stretch in the carotid sinus. cIMT does not closely relate to arterial stiffness until an advanced pathological degree of thickening is reached^33,34^. Early cIMT thickening may not alter stretch of the baroreceptor thus weaking the observed association with HRV explaining our study findings.

A previous study found no association between carotid vascular stiffness indices and total power, LF or HF power in patients with type 2 diabetes^35^ but this could be explained by the failure to account for cardiac autonomic neuropathy (CAN). Others who adjusted for neuropathy in patients with type 2 diabetes did find a significant association with HRV^36^.

It is also possible that, particularly in those with hyperglycaemia, autonomic dysfunction can itself induce arterial stiffness. Parasympathetic dysfunction precedes sympathetic dysfunction, resulting in sympathetic innervation dominating^37^. Indeed, we found that HbA1c–a summary measure of blood glucose levels over the preceding 3 months–was significantly associated with SDNN, HRV triangular index and LF power at univariate analysis and tended towards significance along with HRV triangular index at multivariable analysis, suggesting a potentially significant biological association between HbA1c and HRV. It is also plausible that any link between HRV and elastic arteries reflects changes in their stress/strain relationship (elastance) due to elevated blood sugar and resultant alterations in baroreceptor activation.

Our study did not find an association between previous stroke and HRV in contrast to others^39, 40^ and this is likely due to the small stroke numbers in our cohort. It has been shown that HRV alterations correlate with infarct site, indicating lateralisation of autonomic control. Increased sympathetic discharge may result from injury to the right insular cortex, while parasympathetic increase may be a consequence of left insular cortex damage^41^. Brainstem lesions involving the spinal trigeminal nucleus or rostral ventrolateral medulla may also impact HRV.

Our results demonstrated a significant association between HRV and plasma triglyceride levels at univariate analysis but not with LDL or HDL, replicating findings from another study on non-diabetic individuals^42^ . Authors suggested that the observed association between triglycerides and HRV could be wholly explained by carotid atherosclerotic vascular disease. However, it is also possible that the alteration in autonomic tone could itself also induce hypertriglyceridaemia. This theory is supported by the recognized effect that β blockade with some β-blockers has on serum triglyceride levels^43–46^, likely due to altered sympathetic tone. However, the effect of autonomic tone on lipid metabolism more generally is complex^47^.

We did not find a significant association between physical activity levels and HRV, in contrast to a recent meta-analysis^48^. This discrepancy could be down to the age of our cohort with generally low levels of physical activity being reported. It could be explained by the subjective self-reported physical activity measures used in our study, compared to more objective approaches used by others. Another explanation relates to differences in study design, as some of the interventional studies included in the meta-analysis had prescribed weeks of moderate intensity exercise and measured HRV changes before and after.

### Limitations

Given the cross-sectional study design, it is not possible to imply causality from the observed associations. The inclusion of British people born during the same week in 1946, leads to issues with external validity as the data cannot be easily generalized to non-British populations. Arterial stiffness is highly dependent on blood pressure and we did not pursue recently derived formulae to calculate an arterial stiffness index independent of blood pressure^49^. As already noted, our method for measuring physical activity was subjective.

## Conclusion

Regardless of the presence of carotid atherosclerotic vascular disease (indicated by cIMT), hypertension or stroke, carotid arterial function in older age associates with a dampened HRV response, potentially through an impaired baroreceptor response.

## Data Availability

All data produced are available online at
LHA@ UCL website via skylark

## Data Availability

NSHD data is available from https://www.nshd.mrc.ac.uk/data. Data spreadsheets and statistical codes used for this analysis are provided online in GitHub https://github.com/MaxFornasiero/HRVxCarotidDistensibility/blob/main/Main

## Acknowledgements

The authors would like to thank the NSHD participants for their ongoing engagement with the study and attendance at follow-up for data collection. The authors are also grateful to Imran Shah and Andrew Wong at the MRC Unit for Lifelong Health and Ageing, UCL for facilitating access to the data.

## Author Contributions

All authors contributed significantly to the design, implementation, analysis, interpretation and manuscript writing. The corresponding author attests that all listed authors meet the authorship criteria and that no others meeting the criteria have been omitted.

