## Supplementary Material for "Association between carotid arterial strain and heart rate variability in older age"

a) Institute of Cardiovascular Science, University College London, Gower Street, London WC1E 6BT, UK
b) The Royal Free Hospital, Centre for Inherited Heart Muscle Conditions, Cardiology Department, Pond Street, Hampstead, London NW3 2QG, UK
c) UCL Medical School, Gower Street, London, WC1E 6BT
d) Cardiac MRI Unit, Barts Heart Centre, West Smithfield, London, EC1A 7BE
e) Unit for Lifelong Health and Ageing at UCL, University College London, Fitzrovia, London WC1E 7HB
f) Department of Physiology, Institute of Medicine, Sahlgrenska Academy, University of Gothenburg, Gothenburg, Sweden
g) Swedish Institute for Global Health Transformation (SIGHT), Royal Swedish Academy of Sciences, Stockholm, Sweden.

**Table S1. Missing values per covariate in addition to the complete exposure-outcome pair.**

| **Variable** | **SDNN, n (%)** | **RMSDD, n (%)** | **HRV Triangular Index, n (%)** |
| --- | --- | --- | --- |
| Sex | 0 (0) | 0 (0) | 0 (0) |
| Average CAS | 0 (0) | 0 (0) | 0 (0) |
| SEP at 43yrs | 75 (8.3) | 75 (8.3) | 75 (8.3) |
| BMI | 0 (0) | 0 (0) | 0 (0) |
| Triglyceride | 52 (5.8) | 52 (5.8) | 52 (5.8) |
| HbA1c | 51 (5.7) | 51 (5.7) | 51 (5.7) |
| MI/angina | 70 (7.8) | 70 (7.8) | 70 (7.8) |
| Stroke | 74 (8.3) | 74 (8.3) | 74 (8.3) |
| Hypertension | 0 (0) | 0 (0) | 0 (0) |

Results are reported as counts. Missingness is regarded as significant if >10% and none are present.

*BMI = body mass index; CAS = carotid arterial strain; MI = myocardial infarction; SEP = socioeconomic position; yrs = years.*

**Table S2. Sensitivity analysis showing results of multivariable analysis for the association between CAS and HRV biomarkers after removing participants with known cardiovascular disease**.

| **Variable** |  | **SDNN** | | **RMSDD** | | **HRV triangular index** | |
| --- | --- | --- | --- | --- | --- | --- | --- |
|  | N | *β* Coefficient (95% CI) | *p*-value | *β* Coefficient (95% CI) | *p*-value | *β* Coefficient (95% CI) | *p*-value |
| Age | 846 | -1.70 (-2.6,-0.8) | **<0.001** | -0.33 (-1.2,0.4) | 0.407 | -0.44 (-0.6,-0.3) | **<0.001** |
| Sex | 846 | -2.39 (-4.4,-0.4) | **0.019** | 0.87 (-1.0,2.8) | 0.363 | -0.26 (-0.7,0.2) | 0.231 |
| Average CAS | 846 | 0.43 (0.1,0.7) | **0.003** | 0.49 (0.2,0.8) | **<0.001** | 0.07 (0.0,0.1) | **0.021** |
| SEP at 43yrs | 846 | 0.01 (-2.4,2.6) | 0.993 | -0.18 (-2.5,2.3) | 0.883 | -0.24 (-0.8,0.3) | 0.387 |
| BMI | 846 | -0.13 (-0.4,0.1) | 0.287 | -0.05 (-0.3,0.2) | 0.672 | -0.04 (-0.1,0.0) | 0.165 |
| Triglyceride | 846 | -1.16 (-2.5,0.3) | 0.088 | -1.10 (-2.3,0.2) | 0.076 | -0.23 (-0.5,0.1) | 0.130 |
| HbA1c | 846 | -0.12 (-0.2,0.0) | 0.066 | -0.02 (-0.1,0.1) | 0.782 | -0.03 (-0.1,0.0) | **0.042** |
| Hypertension | 846 | -1.40 (-3.5,0.7) | 0.181 | -0.79 (-2.7,1.2) | 0.424 | -0.26 (-0.7,0.2) | 0.260 |

Significant *p* values are highlighted in bold (*p*<0.05).

*Abbreviations as in* ***Table S1.***

**Table S3. Sensitivity analysis showing results of multivariable analysis for the association between CAS and HRV biomarkers after additional adjustment for cIMT.**

| **Variable** |  | **SDNN** | | **RMSDD** | | **HRV triangular index** | |
| --- | --- | --- | --- | --- | --- | --- | --- |
|  | N | *β* Coefficient (95% CI) | *p*-value | *β* Coefficient (95% CI) | *p*-value | *β* Coefficient (95% CI) | *p*-value |
| Age | 896 | -1.16 (-2.1,-0.3) | **0.013** | -0.16 (-1.0,0.7) | 0.710 | -0.35 (-0.6,-0.2) | **<0.001** |
| Sex | 896 | -2.72 (-4.9,-0.6) | **0.013** | 0.55 (-1.5,2.6) | 0.595 | -0.34 (-0.8,0.1) | 0.152 |
| Average CAS | 896 | 0.50 (0.2,0.8) | **0.001** | 0.55 (0.3,0.9) | **<0.001** | 0.09 (0.0,0.2) | **0.006** |
| SEPat43 | 896 | 0.18 (-2.8,2.6) | 0.894 | -1.86 (-4.2,0.7) | 0.129 | -0.27 (-0.8,0.3) | 0.354 |
| BMI | 896 | -0.07 (-0.3,0.2) | 0.617 | 0.03 (-0.2,0.3) | 0.833 | -0.02 (-0.1,0.0) | 0.470 |
| Triglyceride | 896 | -0.96 (-2.4,0.6) | 0.203 | -1.09 (-2.4,0.4) | 0.110 | -0.19 (-0.5,0.1) | 0.245 |
| HbA1c | 896 | -0.11 (-0.2,0.0) | 0.114 | -0.05 (-0.2,0.1) | 0.443 | -0.03 (-0.1,0.0) | 0.056 |
| MI/angina | 896 | -2.49 (-6.9,2.7) | 0.298 | 3.13 (-1.8,9.4) | 0.250 | -0.38 (-1.4,0.8) | 0.479 |
| Stroke | 896 | -0.87 (-1.8,0.7) | 0.153 | -0.63 (-1.4,1.2) | 0.257 | -0.14 (-0.4,0.2) | 0.350 |
| Hypertension | 896 | -1.63 (-3.8,0.6) | 0.149 | -1.59 (-3.7,0.6) | 0.137 | -0.34 (-0.8,0.2) | 0.175 |
| cIMT | 896 | 3.00 (-6.0,12.4) | 0.506 | 6.66 (-1.8,15.7) | 0.123 | 0.36 (1.5,2.3) | 0.716 |

Significant *p* values are highlighted in bold (*p*<0.05). cIMT = carotid intima-media thickness. Other abbreviations are as in **Table S1.**

**Table S4. Univariate analysis results for the remaining HRV variables: total PSD, normalised LF power, PSD squared and LF/HF ratio.**

| **Variable** | **Total PSD** | | | **Normalised LF power** | | | **PSD squared** | | | **LF/HF ratio** | |
| --- | --- | --- | --- | --- | --- | --- | --- | --- | --- | --- | --- |
|  | ***β* coefficient (95% CI)** | ***p*-value** | ***β* coefficient (95% CI)** | | ***p*-value** | ***β* coefficient (95% CI)** | | ***p*-value** | ***β* coefficient (95% CI)** | | ***p*-value** |
| **Carotid Variables** | | | | | | | | | | | |
| Cross-sectional CAS left | 20.68 (6.5,36.1) | **0.010** | -0.22 (-0.50,0.07) | | 0.144 | 0.00 (0.00,0.00) | | **0.050** | -0.01 (-0.04,0.03) | | 0.702 |
| Cross-sectional CAS right | 14.88 (1.9,28.6) | **0.047** | -0.24 (-0.52,0.04) | | 0.099 | 0.00 (0.00,0.00) | | 0.123 | -0.01 (-0.04,0.02) | | 0.629 |
| Average CAS | 21.41 (6.3,37.6) | **0.013** | -0.28 (-0.59,0.03) | | 0.088 | 0.00 (0.00,0.00) | | **0.044** | -0.01 (-0.04,0.03) | | 0.651 |
| Average cIMT | 187.40 (-307.7,707.9) | 0.460 | -1.09  (-10.80,8.85) | | 0.829 | 0.13  (-0.04,0.06) | | 0.604 | -0.22 (-1.41,1.04) | | 0.728 |
| IMT maximum | 177.00 (-229.0,606.3) | 0.398 | -0.71 (-8.64,7.46) | | 0.865 | 0.01  (-0.03,0.05) | | 0.710 | -0.15 (-1.12,0.89) | | 0.767 |
| **Demographics** | | | | | | | | | | | |
| Age | -67.50 (-118.9,-19.7) | **0.010** | -0.50 (-1.56,0.54) | | 0.357 | 0.01 (0.00,0.01) | | **0.013** | 0.03 (-0.10,0.16) | | 0.618 |
| Male | -91.85 (-214.5,27.6) | 0.135 | -6.96 (-9.42,-4.52) | | **<0.001** | 0.00  (-0.01,0.02) | | 0.472 | -0.64 (-0.97,-0.32) | | **<0.001** |
| Social class 1989: | -17.00 (-159.2,144.7) | 0.825 | -2.57 (-5.63,0.61) | | 0.106 | 0.00  (-0.02,0.01) | | 0.712 | -0.25 (-0.62,0.17) | | 0.214 |
| **Anthropometrics** | | | | | | | | | | | |
| BMI | -5.71 (-18.6,8.3) | 0.405 | -0.34 (-0.62,-0.05) | | **0.014** | 0.00 (0.00,0.00) | | 0.842 | -0.03 (-0.06,0.01) | | 0.119 |
| Waist-to-hip ratio | -169.90 (-925.6,597.9) | 0.657 | 17.64 (2.42,32.86) | | **0.023** | 0.02  (-0.06,0.09) | | 0.623 | 1.17 (-0.77,3.11) | | 0.237 |
| **Cardiac** | | | | | | | | | | | |
| Mean DBP | -0.44 (-6.5,5.7) | 0.888 | 0.19 (0.06,0.32) | | **0.003** | 0.00 (0.00,0.00) | | **0.034** | 0.01 (0,0.03) | | 0.071 |
| Mean SBP | -0.35 (-3.6,3.0) | 0.835 | 0.06 (-0.01,0.13) | | 0.069 | 0.00 (0.00,0.00) | | **0.042** | 0.01 (0,0.01) | | 0.190 |
| LV mass | 0.19 (-0.7,1.2) | 0.678 | 0.00 (-0.02,0.02) | | 0.715 | 0.00 (0.00,0.00) | | 0.272 | 0 (0,0) | | 0.440 |
| Ejection Fraction bi-plane | 0.98 (-7.6,9.1) | 0.825 | -0.98 (-0.27,0.08) | | 0.284 | 0.00 (0.00,0.00) | | 0.253 | -0.01  (-0.03,0.01) | | 0.436 |
| **Blood Markers** | | | | | | | | | | | |
| Total Cholesterol | -15.29 (-68.2,39.2) | 0.572 | 0.37 (-0.75,1.48) | | 0.510 | 0.00 (0.00,0.01) | | 0.847 | 0.07 (-0.07,0.21) | | 0.325 |
| HDL ratio | -7.94 (-75.9,64.6) | 0.813 | 0.76 (-0.60,2.14) | | 0.271 | 0.00  (-0.01,0.01) | | 0.860 | 0.08 (-0.09,0.27) | | 0.355 |
| LDL | 1.29 (-61.7,66.2) | 0.968 | 0.41 (-0.89,1.72) | | 0.529 | 0.00  (-0.01,0.01) | | 0.958 | 0.07 (-0.1,0.24) | | 0.420 |
| Triglyceride | -78.09 (-105.5,-13.3) | **0.004** | 0.77 (-0.84,2.52) | | 0.366 | 0.00  (-0.01,0.01) | | 0.930 | 0.1 (-0.1,0.34) | | 0.358 |
| HbA1c | -9.53 (-12.4,-4.0) | **<0.001** | -0.07 (-0.23,0.11) | | 0.448 | 0.00 (0.00,0.00) | | 0.741 | -0.01  (-0.02,0.01) | | 0.580 |
| **Other Clinical Factors** | | | | | | | | | | | |
| Smoking | -46.38 (-141.4,42.1) | 0.353 | 1.08 (-0.93,3.04) | | 0.281 | 0.00  (-0.01,0.01) | | 0.448 | 0.27 (0.02,0.5) | | **0.024** |
| Exercise levels | -0.11 (-0.5,0.4) | 0.624 | -0.02 (-0.03,-0.01) | | **<0.001** | 0.00 (0.00,0.00) | | 0.561 | 0 (0,0) | | **<0.001** |
| MI or angina | -151.70 (-358.5,136.1) | 0.216 | -3.88 (-9.00,1.81) | | 0.158 | -0.02  (-0.04,0.01) | | 0.147 | -0.63  (-1.12,0.02) | | **0.024** |
| Stroke | -66.30 (-88.7,-5.4) | **<0.001** | 0.38 (-1.15,2.47) | | 0.681 | 0.00  (-0.01,0.01) | | 0.902 | -0.01  (-0.14,0.34) | | 0.927 |
| Diabetes mellitus | 131.16 (-389.2,144.0) | 0.187 | -1.63 (-7.42,4.93) | | 0.604 | 0.01  (-0.02,0.04) | | 0.505 | 2.56 (-0.56,1.18) | | 0.742 |
| Hypertension, n (%) | -46.57 (-166.6,76.7) | 0.451 | 2.20 (-0.29,4.71) | | 0.085 | 0.01 (0.00,0.02) | | 0.215 | 0.21 (-0.1,0.54) | | 0.196 |
| Hypercholesterolaemia, n (%) | -6.38 (-196.1,235.8) | 0.953 | -2.43 (-6.48,1.94) | | 0.257 | -0.02  (-0.03,0.01) | | 0.122 | -0.31  (-0.76,0.25) | | 0.219 |
| History of cardiovascular event, n (%) | -108.23 (-317.6,182.6) | 0.383 | -4.47 (-9.36,0.95) | | 0.088 | -0.01  (-0.03,0.02) | | 0.441 | -0.67  (-1.34,-0.04) | | **0.013** |

Significant p values are highlighted in bold (p<0.05).

DBP = diastolic blood pressure. SBP = systolic blood pressure. HDL = high-density lipoprotein. LDL = low-density lipoprotein. LV = left ventricular. HF = high-frequency. LF = low-frequency. Other abbreviations as in **Tables S1** and **S2.**

**Table S5. Multivariable analysis for the association of CAS with the remaining HRV biomarkers: PSD squared and total PSD.**

| **Variable** | **Power Spectral Density Squared** | | **Total Power Spectral Density** | |
| --- | --- | --- | --- | --- |
|  | *β* Coefficient (95% CI) | *p*-value | *β* Coefficient (95% CI) | *p*-value |
| Age | 0.01 (0.0,0.0) | **0.001** | -42.92 (-101.7,15.9) | 0.152 |
| Sex | 0.00 (0.0,0.0) | 0.587 | -147.81 (-284.6,-11.0) | **0.034** |
| Average CAS | 0.00 (0.0,0.0) | 0.671 | 19.01(-0.1,38.1) | 0.051 |
| Social Class 1989 | 0.00 (0.0,0.0) | 0.746 | -28.20 (-196.1,139.7) | 0.742 |
| BMI | 0.00 (0.0,0.0) | 0.608 | 0.45 (-16.4,17.3) | 0.959 |
| Triglyceride | 0.00 (0.0,0.0) | 0.828 | -39.90 (-130.1,50.3) | 0.386 |
| HbA1c | 0.00 (0.0,0.0) | 0.123 | -6.14 (-12.8,0.6) | 0.073 |
| MI/angina | -0.01 (0.0,0.0) | 0.660 | -11.31 (-343.5,320.9) | 0.947 |
| Stroke | -0.01 (0.0,0.0) | 0.222 | -42.82 (-99.1,13.4) | 0.135 |
| Hypertension | 0.02 (0.0,0.0) | **0.036** | -104.99 (-244.8,34.8) | 0.141 |

Significant p values are highlighted in bold (p<0.05). Abbreviations as in **Table 1.**
